# Estimation and correlation of sirtuin1 with Carboxy Methyl Lysine in type 2 diabetic and its microvascular complication

**DOI:** 10.1101/2020.07.27.20163253

**Authors:** Sai Deepika Ram Mohan, Shashidhar Kurpad Nagaraj, Raveesha Anjanappa, Muninarayana Chandrappa

## Abstract

**Background:** Increase in Diabetes Mellitus increases incidence of its Microvascular complications such as diabetic retinopathy, diabetic kidney disease (DKD), neuropathy, stroke and cardio vascular diseases (CVD). Advanced glycation end products promotes Type 2 Diabetes to its major Microvascular complication; diabetic kidney disease or diabetic nephropathy leading to increase in expression of sirtuin1; a regulatory protein mediating deacetylation of histone proteins. In addition to diet and nutrition, environmental changes may increase incidence of disorders, one such factor considered in this study is Fluoride.

**Objective:** Estimation of Sirtuin1 in type 2 diabetes mellitus and diabetic kidney disease and interpreting the outcome by diabetic profiling of patients with controls. Participants: 150 study subjects were recruited for this cross-sectional study divided into 3groups with 50 subjects in each group.

**Methods:** Diabetic and renal profiling was carried by fully automated analyzer available in our hospital facility, eGFR was calculated, sirtuin1 and CML were measured by ELISA, serum and urine fluoride were estimated by Ion Selective Electrode.

**Results:** Significant differences observed in FBS, PPBS and CML of deceased with controls. Least median of sirt1 was observed in diabetic nephropathy (36.9). Also, urine and serum fluoride levels were proportionally balanced in group 1& 2 in contrast with group 3 [0.28 (0.2-0.54) & 0.2 (0.15-0.26)].

**Conclusion:** Decrease in Sirtuin1 in group 3 may be due to chronic hyperglycemia and oxidative stress in diabetes hence, further research on large cohort may aid considering sirtuin1 as a biomarker or therapeutic target in aging disorders.

## Introduction

Quality of day to day living and livelihood of human being is influenced by ready available packed food so called fast food, due to rapid urbanization across the globe^1^. Lifestyle matters for body organs and cells to follow their routine for synthesis and secretion of biochemicals for bodily metabolism^2^. Lack of proper nutrients in food results in various aging disorders, majorly booming in health sector in recent days are cancer, metabolic syndrome and diabetes mellitus. Based on reports by World Health Organization (WHO), diabetes mellitus (DM) and its Microvascular complication will be one of the causes for increase in morbidity and mortality by 2030^3^. India being heading the list of highest diabetes cases will continue to be so till 2030 with about 2.5 folds increase in incidence compared to the year 2000^3^.

Type 2 diabetes mellitus (T2DM) is most common which is either due to improper secretion of insulin by pancreas or by improper utilization of secreted insulin by peripheral cells^4^. Due to increase in prevalence of T2DM there is an increased incidence of its Microvascular complications such as diabetic retinopathy, diabetic kidney disease (DKD), diabetic neuropathy, stroke and cardio vascular diseases (CVD)^5,6^. All of these diabetic microvascular complications are aging disorder since they are resultant of cell stress generated during hyperglycaemia leading to formation of free radicals which in turn forms a group of chemicals called reactive oxygen species (ROS)^7^. Increase in ROS increases rate of apoptosis (programmed cell death) causing imbalance in proportion of cell death and cell division. During the imbalance, expression of an enzyme called ‘Sirtuin1 (Sirt1)’ increases which initiates Autophagy of damaged cells wherever necessary^6^. Fyre in 1999 discovered 5 human homologues sirt1-5 similar to yeast (Saccharomyces. cerevisiae) sir2 (Silent Information Regulator 2) gene, one among them is Sirt1. Sirt1 is a NAD+ dependant deacetylase enzyme, deacetylating histones, transcriptional factors etc^9-11^.

There are several other mechanisms contributing to increasing incidents of vascular damage in hyper glycaemic called advanced glycation end products (AGE). AGE are produced non-enzymatically as a part of body metabolism and also acquired through diet^12^. Since AGE promotes T2DM to its major Microvascular complication especially diabetic kidney disease or diabetic nephropathy, it leads to increase in expression of sirtuin1^13^. One such significant AGE in research in present era is Carboxy methyl lysine (CML) which is formed either due to improper glucose metabolism and/or externally through diet^14^. The majority of CML in tissue is derived from lipid peroxidation reactions, even during hyperglycemias when concentrations of glucose and Amadori products on protein are increased^15^.The mechanism of formation of CML and other AGEs during carbohydrate oxidation reactions is still uncertain^16^. In addition to diet and nutrition environmental changes also has scope in increasing incidence of disorders, one such factor considered in this study is Fluoride (F).

Chronic F exposure leads to various types of Fluorosis such as dental fluorosis, skeletal fluorosis and recent studies are carried out on non-skeletal fluorosis. There are various animal studies on non-skeletal fluorosis concluding that deposition of F in blood vessels and organs leads to various complication in F endemic areas^17^. Since Kolar is considered F endemic area due to increased water F level of about 0.6-4ppm in villages around Kolar district^18^. After metabolism, excretion of F is mainly through renal tissue, partly through faeces and sweat^17^. Therefore, decrease in F clearance indicates decreased filtering capacity of kidneys which is estimated in this study for comparison between controls, T2DM and Diabetic Kidney Disease.

## Methods

Study subjects with advice of physician were recruited from diabetic outpatient clinic of the hospital attached to the college where the research was carried out. A total of 150 study subjects were divided into 3 groups with 50 subjects in each group, study design being cross-sectional approved by Central Ethics Committee (CEC) of the academy a registered body under Indian government. DKD patients under dialysis, CKD due to hypertension, nephritis, sepsis, were excluded. All subjects included in this study are residents of Kolar district for minimum of 5years consuming filtered bore well water. Informed consent was obtained from all the study subjects complying declaration of Helsinki which was also approved by CEC.

Patients and controls were instructed to fast for a minimum of 8hour for investigating blood parameters which include fasting blood glucose (FBS), post prandial blood glucose (PPBS), blood urea, serum creatinine, serum uric acid, sodium, potassium analyzed by fully automated Vitros 5, 1 FS glycosylated hemoglobin (HbA1c) estimated by HPLC using BioRad D10, eGFR was calculated according to KDQIO 2012 guidelines and anthropometric parameters and demographic data were also recorded. Written informed consent was obtained from study subjects approved by IEC. Sirt1 was measured by sandwich ELISA (kits procured from Sincere Biotech China). Serum and Urine fluoride were estimated by Orion Thermo Scientific Fluoride Ion Selective Electrode (ISE).

## Statistical analysis

SPSS version 22 by IBM was used for data analysis. Routine parameters were represented as Mean ± S.D sirtuin1 is presented as median (interquiles). Pearson’s correlation and ANOVA test were used to calculate significance for normally distributed data. Spearman’s correlation, Kruskal-walli’s test were applied to calculate p-value of non-normally distributed variables. p-Value < 0.05 was considered significant.

## Results

Age and gender matched subjects were recruited. Female subjects recruited were 73 in number and the remaining 77 were males. Most of the subjects in group 2 that is, diabetic nephropathy were males (n= 32). From table1 and table 2, since, T2DM complications arise during latent stages of disorder, hence, age is significant with p-value 0.000 between group 1&2 and group 1&3. Mild increase in blood pressure was seen between group 1 and 2, major significant difference was observed between group1 & 3 and group 2 & 3 which is evident that DKD cases are hypertensive. During consenting procedure it was also noticed that 50% of group 2 subjects were consuming directly the groundwater for drinking and cooking unlike the other groups who were using filtered water for drinking and bore well water for cooking.

**Table 1:**
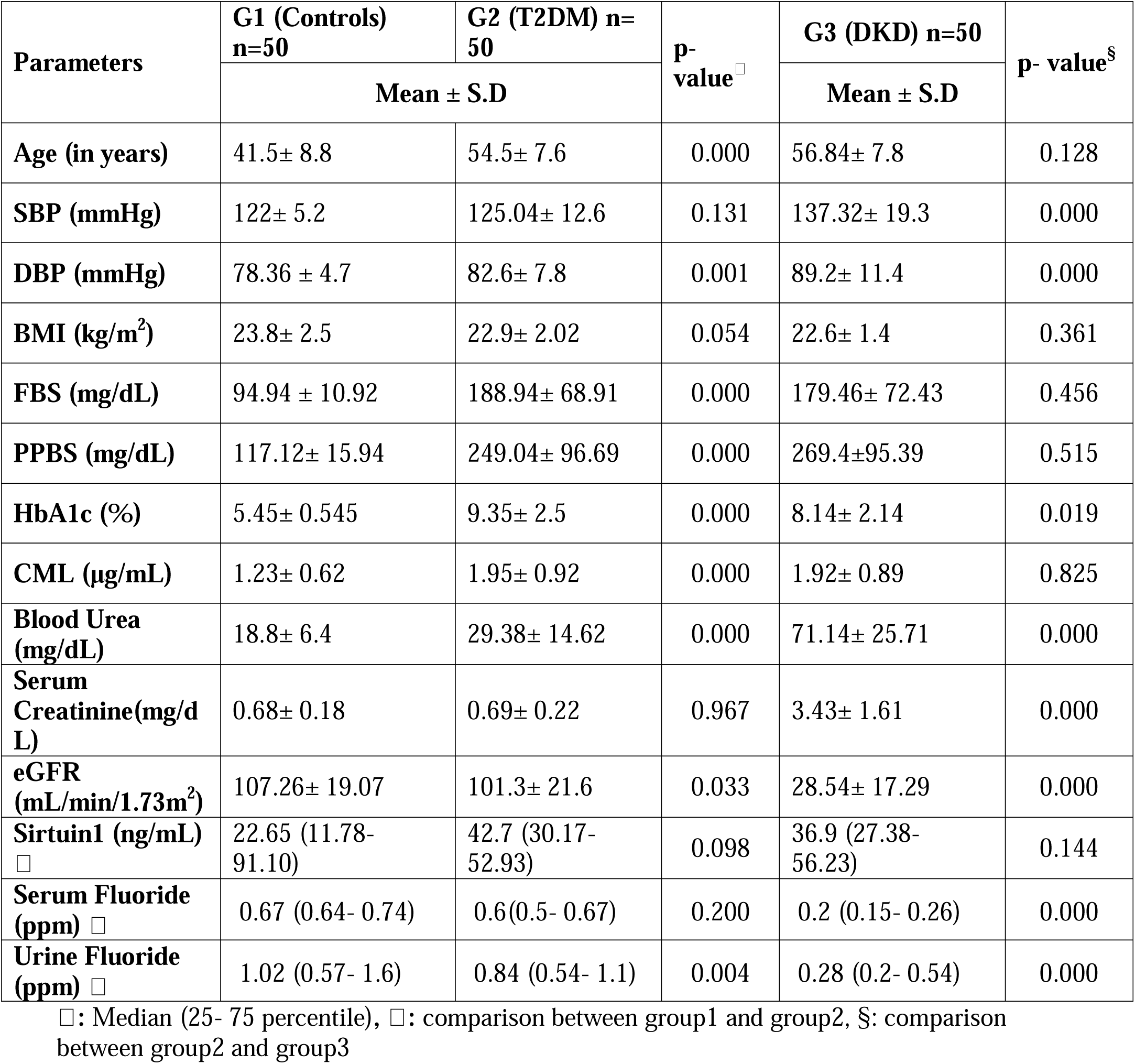
Comparison of Demographic data, diabetic and renal profile

| Parameters | G1 (Controls)<br>n=50 | G2 (T2DM) n=50 | p-value<sup>□</sup> | G3 (DKD) n=50 | p- value<sup>§</sup> |
| --- | --- | --- | --- | --- | --- |
|  | Mean ± S.D |  |  | Mean ± S.D |  |
| Age (in years) | 41.5± 8.8 | 54.5± 7.6 | 0.000 | 56.84± 7.8 | 0.128 |
| SBP (mmHg) | 122± 5.2 | 125.04± 12.6 | 0.131 | 137.32± 19.3 | 0.000 |
| DBP (mmHg) | 78.36 ± 4.7 | 82.6± 7.8 | 0.001 | 89.2± 11.4 | 0.000 |
| BMI (kg/m<sup>2</sup>) | 23.8± 2.5 | 22.9± 2.02 | 0.054 | 22.6± 1.4 | 0.361 |
| FBS (mg/dL) | 94.94 ± 10.92 | 188.94± 68.91 | 0.000 | 179.46± 72.43 | 0.456 |
| PPBS (mg/dL) | 117.12± 15.94 | 249.04± 96.69 | 0.000 | 269.4±95.39 | 0.515 |
| HbA1c (%) | 5.45± 0.545 | 9.35± 2.5 | 0.000 | 8.14± 2.14 | 0.019 |
| CML (µg/mL) | 1.23± 0.62 | 1.95± 0.92 | 0.000 | 1.92± 0.89 | 0.825 |
| Blood Urea (mg/dL) | 18.8± 6.4 | 29.38± 14.62 | 0.000 | 71.14± 25.71 | 0.000 |
| Serum Creatinine(mg/d L) | 0.68± 0.18 | 0.69± 0.22 | 0.967 | 3.43± 1.61 | 0.000 |
| eGFR (mL/min/1.73m<sup>2</sup>) | 107.26± 19.07 | 101.3± 21.6 | 0.033 | 28.54± 17.29 | 0.000 |
| Sirtuin1 (ng/mL)<br>□ | 22.65 (11.78-91.10) | 42.7 (30.17-52.93) | 0.098 | 36.9 (27.38-56.23) | 0.144 |
| Serum Fluoride (ppm)<br>□ | 0.67 (0.64- 0.74) | 0.6(0.5- 0.67) | 0.200 | 0.2 (0.15- 0.26) | 0.000 |
| Urine Fluoride (ppm)<br>□ | 1.02 (0.57- 1.6) | 0.84 (0.54- 1.1) | 0.004 | 0.28 (0.2- 0.54) | 0.000 |

□: Median (25- 75 percentile), □: comparison between group1 and group2, §: comparison between group2 and group3

**Table 2:**
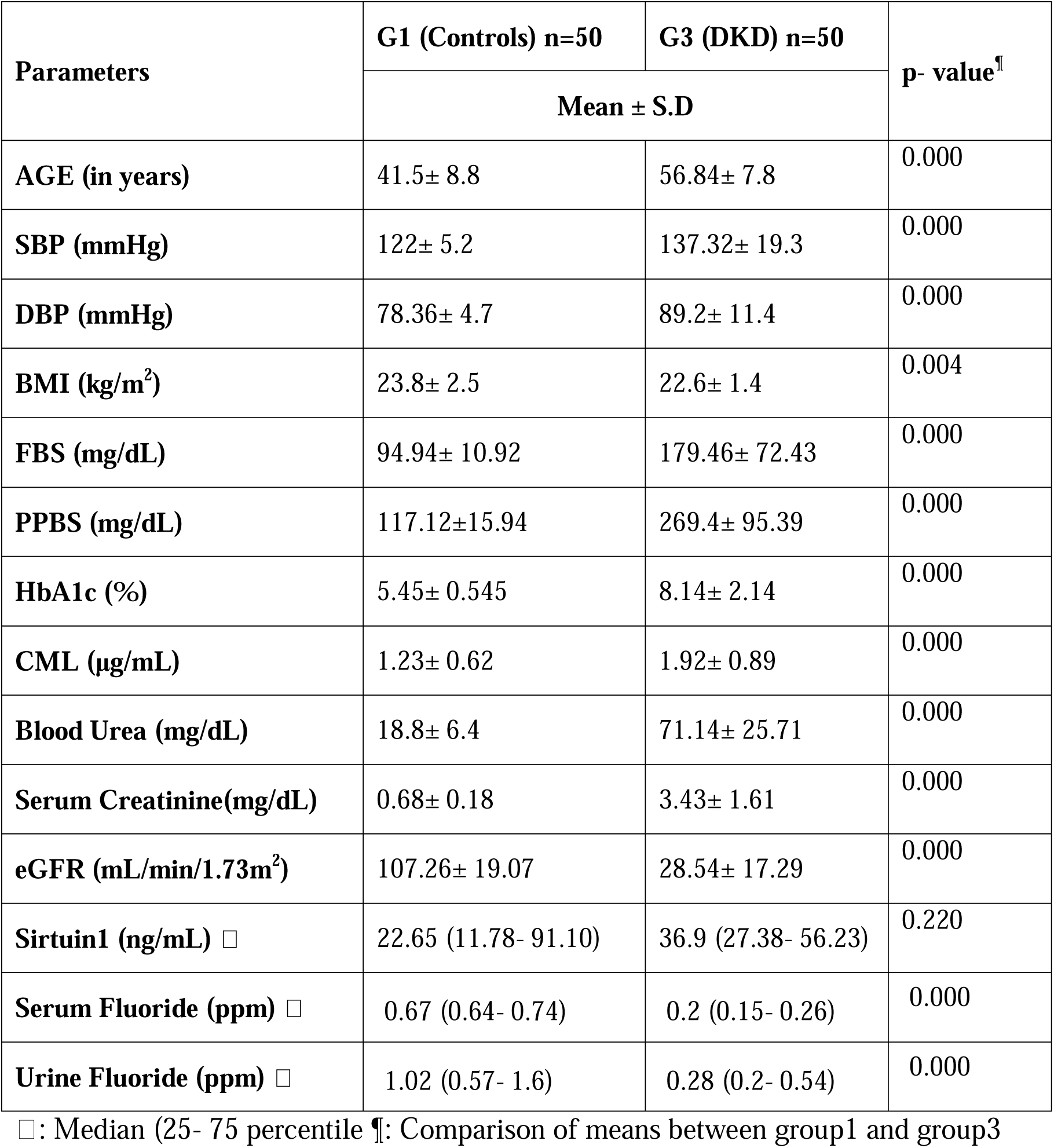
Comparison of Demographic data, diabetic and renal profile between group1 and group3 subjects

| Parameters | G1 (Controls) n=50 | G3 (DKD) n=50 | p- value<sup>¶</sup> |
| --- | --- | --- | --- |
|  | Mean ± S.D |  |  |
| AGE (in years) | 41.5± 8.8 | 56.84± 7.8 | 0.000 |
| SBP (mmHg) | 122± 5.2 | 137.32± 19.3 | 0.000 |
| DBP (mmHg) | 78.36± 4.7 | 89.2± 11.4 | 0.000 |
| BMI (kg/m<sup>2</sup>) | 23.8± 2.5 | 22.6± 1.4 | 0.004 |
| FBS (mg/dL) | 94.94± 10.92 | 179.46± 72.43 | 0.000 |
| PPBS (mg/dL) | 117.12±15.94 | 269.4± 95.39 | 0.000 |
| HbA1c (%) | 5.45± 0.545 | 8.14± 2.14 | 0.000 |
| CML (µg/mL) | 1.23± 0.62 | 1.92± 0.89 | 0.000 |
| Blood Urea (mg/dL) | 18.8± 6.4 | 71.14± 25.71 | 0.000 |
| Serum Creatinine(mg/dL) | 0.68± 0.18 | 3.43± 1.61 | 0.000 |
| eGFR (mL/min/1.73m<sup>2</sup>) | 107.26± 19.07 | 28.54± 17.29 | 0.000 |
| Sirtuin1 (ng/mL) □ | 22.65 (11.78- 91.10) | 36.9 (27.38- 56.23) | 0.220 |
| Serum Fluoride (ppm) □ | 0.67 (0.64- 0.74) | 0.2 (0.15- 0.26) | 0.000 |
| Urine Fluoride (ppm) □ | 1.02 (0.57- 1.6) | 0.28 (0.2- 0.54) | 0.000 |

□: Median (25- 75 percentile) ¶: Comparison of means between group1 and group3

Significant differences were observed in mean of FBS, PPBS, HbA1c and CML of Group 2 & 3 with group 1. Major significant difference was seen in renal profile parameters; blood urea, serum creatinine and eGFR. Blood urea and serum creatinine was increased compared to control and diabetic group matching ADA criteria of T2DM and DKD. In contrast, estimated Glomerular filtration rate was drastically decreased and was <30 (28.54 ± 17.29) as per KDIGO guidelines^**13**^. Sirtuin1 and Fluoride are represented as median (25-75 percentile) since it has a broad range hence not normally distributed. Least median range was observed in group1 [22.65 (11.78-91.10)], projected median for group3 is 36.9 (27.38-56.23) which is also lesser than group 2 [42.7 (30.17-52.93)]. Also, there was a significant difference of serum and urine F between groups.

Table 3 depicts Spearmann’s correlation (ρ) analysis explaining the trend in of a parameter in comparison with the special molecule sirtuin1. Sirt1 is correlated with AGE; CML and serum fluoride, showing a negative trend, indicating protective effect of increase in sirt1 may decrease formation of CML. This parameter is first time compared with sirtuin1 in human population hence, further invitro studies are required to study in detail the effects of sirtuin1 on different tissues and correlate in different conditions.

**Table 3:**
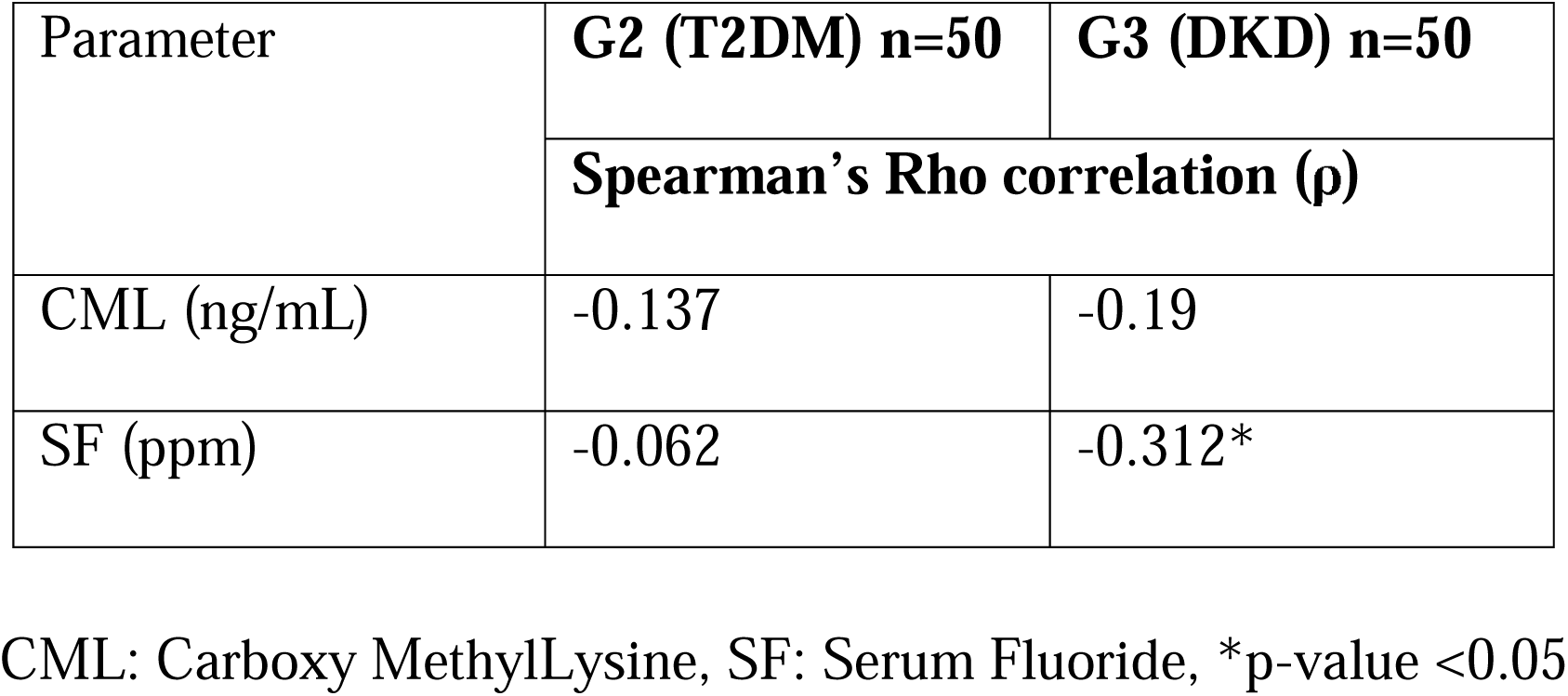
Correlation of Sirtui1 with CML and serum fluoride.

## Discussion

There are several animal studies involving sirtuin1 in pathological conditions such as, T2DM, cancer, metabolic syndrome, mineral toxicity etc. Also, there are invitro studies which include sirtuin1 for understanding pathogenesis and signalling mechanisms for sirtuin1 metabolism. All these studies forms the basis for comparison and correlation of blood parameters in T2DM and its Microvascular complication; diabetic kidney disease (DKD) included in this study.

DKD is categorized based on KGIDO 2012 guidelines to consider either Albuminuria or eGFR. This study calculated eGFR involving equation with only serum creatinine and eliminated those cases whose eGFR is mildly to severly decrease falling under 3rd and 4th stage of DKD with eGFR ranging 25-44 mL/min/1.73m^2 13^. Patients under dialysis are include in 5th stage; renal failure who were excluded from the study.

Sirtuin1 is the major regulator of key transcriptional factors responsible for protein turn over and metabolism^14^. There are no many studies regarding sirtuins and insulin sensitivity in human who may be due to confounding factors such as genetics, environment, dietary habits etc. which leads to broad range of sirtuin values in human. Sirtuins are said to increase during defensive mechanisms wherever possible and necessary. Since, sirtuin1 is localized in nucleus, severe damage of cells at the level of nucleus may decrease synthesis of sirtuin1 as a result and defensive mechanism of sirt1 against oxidative stress and other aging mechanisms are hindered and left irreparable. Apart from ROS generation, advanced glycation end (AGE) products also increase oxidative damage to cells speeding up rate of Microvascular complications in T2DM^15^. Effat A. Khowailed et al in animal model proved that regular metformin administration to T2DM suppresses the pathophysiology which decreases sirt1 and improvement in sirt1 levels were observed^16^. Controlled Hyperglycemia will prevent consequences of diabetes and improve status of all protective molecules in body one such example being Sirtuin1.

Sirt1 involves in Autophagy which is promoted in damaged cells, hence retrieving normal functioning of tissues. Recent researchers are working on sirt1 as therapeutic target in various aging disorders thereby preventing further complications to tissues. Recently it is proved that, dietary restriction improves sirtuin1 levels and decreases AGE especially CML, which is one of the crucial points to be considered in management of T2DM in preventing further complications^19,20^. Interventional studies involving molecules which stimulate sirtuin1 in turn protecting cells from oxidative damage will help improving therapeutics naturally and preventing consequences of disorders. One such molecule now in research is resveratrol which not only increase sirt1 but also improves various other metabolisms in body preventing oxidative stress and ROS generation^21^.

There are very limited studies emphasizing on effects of fluoride on human cells and tissues since its effects are still in invitro state. This study is conducted as preliminary step to elucidate and find effect of circulating F and renal clearance hence estimated serum and urine fluoride (F). Animal study by Takamasa Kido et al. revealed damage to renal tissues in chronic exposure to F^22^. Water fluoride exposure may be the main cause of F exposure in localites of this study, since kidney is the major route of excretion renal damage is expected which is evident from proportionate decrease of urine F correlated with serum F^23^. Table 3 shows negative correlation of serum Sirt1 with CML and serum fluoride in all groups 2 & 3 indicating, increase in Sirt1 may help protecting CML hence can be tried in therapeutics. There is a critical observation made with respect to the graph that, off late, sirtuin1 is being targeted for therapeutic uses in chronic aging related illness. Therefore, taking in account the sirtuin1 as a regulatory protein may help research to administer sirtuin1 and analyse the process of aging and evaluate its counter action as anti-aging molecule thus preventing chronic illness.

Major drawback of the study is water fluoride estimations of villages were not performed which might have given a clear picture regarding exposure and outcome. Also, comparison of parameters between F exposed and non-exposed population may give a clear picture of the scenario. Molecular study of situin1 one could have explained correlation appropriately.

## Conclusion

Decrease in sirtuin1 levels was observed in group1 and 3 compared to group2 may be due to least damage to cells since pathological exposure to Reactive Oxygen Species generated during oxidative stress in negligible group1. On the other hand, severe damage to cells and chronic hyperglycemia may be the reason for decreased synthesis of sirtuin1 thereby incompatibility of the body to prevent further damage to cells of kidney. Increase of sirt1 in group2 is probably due to protective action of cells from further damage in hyperglycemia and AGE or ROS insult to cells,

Other important molecule of the study Fluoride (F), may not be cause for diabetes but may accelerate diabetes to progress to its microvascular complications as resulted in this study Hence, sirtuin1 is considered as a biomarker for aging disorders especially diabetes and also preventive molecule therefore, introducing Sirtuin1 in therapeutics may help combat the imbalance of cellular damage and repair mechanism.

## Data Availability

All the subjects included in this study had written informed consent according to the guidelines of declaration of Helsinki and the samples and data are stored anonymously for future reference if any

## Acknowledgment

We sincerely thank our academy for encouraging and providing us the facility to select, collect and analyse the sample at our ease.

